# Development of a Multi-Model Ensemble Tool for Early Prediction of 48-Hour Respiratory Failure Risk in CAP Patients

**DOI:** 10.1101/2025.11.06.25339737

**Authors:** Yali Xu, Shubin Guo, Ying Chen, Andong Li, Qi Lyu, Xue Mei

**Author notes:** Corresponding Author: Shubin Guo^a^,Prof; Xue Mei^a^,Prof; Department of Emergency Medicine Clinical Research Center Beijing Chaoyang Hospital Affiliated to Capital Medical University No. 8,Gongrentiyuchang South Road,Chaoyang District, Beijing,100020 China.

## Abstract

**Objective:** To develop a predictive tool capable of early identification of the risk of acute respiratory failure within 48 hours of hospital admission in patients with community-acquired pneumonia (CAP).

**Method:** A retrospective cohort of 257 CAP patients (median age: 76.0 years, IQR: 68.0–84.0; 56.4% male) was analyzed, among whom 148 (57.6%) developed respiratory failure within 48 hours. From 55 clinical variables, key predictors were selected using LASSO regression. Predictive models were then constructed using multivariable logistic regression (MLR) and machine learning algorithms including XGBoost, LightGBM, and Random Forest. To address the probability calibration issue of the XGBoost model, Platt scaling was applied. A final ensemble model was built by weighted averaging of the calibrated XGBoost, LightGBM, and MLR models. Feature importance was analyzed using SHAP (SHapley Additive exPlanations), and clinical utility was evaluated via decision curve analysis (DCA) and calibration plots.

**Result:** Respiratory rate, TNF-α, IL-1β, heart rate, pleural effusion, and body temperature were identified as the most important predictors. Other key features included total bilirubin, serum calcium, albumin/globulin ratio, and platelet count. The weighted ensemble model outperformed individual models, achieving an AUC of 0.792 on the test set.

**Conclusion:** We developed a predictive tool based on multi-model ensemble learning and interpretable machine learning techniques (SHAP), which provides a basis for early risk stratification and prevention of acute respiratory failure in hospitalized CAP patients.

## 1. Introduction

Community-acquired pneumonia (CAP) is defined as an acute infection of the lower respiratory tract involving the lung parenchyma that is acquired outside of hospital settings. It remains one of the top five causes of mortality worldwide [1]. Despite ongoing advances in diagnostic technologies and antimicrobial therapies, a subset of CAP patients experience rapid disease progression after hospital admission, leading to the development of acute respiratory failure (ARF). This complication significantly complicates clinical management and markedly increases mortality risk. As one of the most common and severe complications of CAP, ARF is a major driver of poor clinical outcomes in these patients [2]. CAP patients who develop ARF are substantially more likely to require mechanical ventilation [3], with reported mortality rates reaching up to 50% [4]. Even among survivors, long-term adverse effects may persist. Hypoxemia associated with respiratory failure increases the risk of neurodegeneration, cognitive impairment [5], and delirium [6] in elderly patients, leading to reduced quality of life post-discharge [7]. Therefore, the early identification of CAP patients at risk of developing ARF during hospitalization is of critical clinical importance.

Machine learning (ML) techniques have emerged as powerful tools for analyzing diverse and complex data types, and they are increasingly being applied in predictive healthcare analytics. While traditional risk stratification tools such as the CURB-65 score and the Pneumonia Severity Index (PSI) are widely recommended for CAP assessment, phenotype-based analyses suggest that these scoring systems rely on a limited number of clinical variables and may fail to capture the heterogeneity present in multimodal clinical data [4]. As a result, these conventional tools may fall short in accurately identifying CAP patients at high risk for ARF, limiting their utility in precision early warning systems. In contrast, ML algorithms are well suited to analyze high-dimensional and complex clinical datasets, as they can capture both linear and nonlinear interactions among variables, thereby uncovering synergistic relationships within the data [8]. ML-based approaches have shown significant promise in predicting respiratory failure [9]; however, studies focusing specifically on the early prediction of ARF in CAP patients remain scarce.

In this study, we developed clinical prediction models to assess the risk of ARF within the first 48 hours of hospitalization in patients with CAP. We employed logistic regression and five ML algorithms: Extreme Gradient Boosting (XGBoost), Light Gradient Boosting Machine (LightGBM), k-Nearest Neighbors (KNN), Support Vector Machine (SVM), and Random Forest. These models were trained on clinical data collected at the time of hospital admission. To enhance robustness and minimize the bias associated with any single algorithm, we combined the three best-performing models using a weighted ensemble approach. Finally, to improve model interpretability, we utilized SHapley Additive exPlanations (SHAP) to identify and rank the most important predictive features.

## 2. Materials and Methods

### 2.1 Study Design and Population

This was a retrospective observational study. The study population consisted of patients admitted to the Emergency Department of Beijing Chaoyang Hospital for community-acquired pneumonia (CAP) and subsequently hospitalized between September 2024 and March 2025.Inclusion criteria were as follows:1. Age ≥18 years; 2. Diagnosis consistent with CAP, defined as radiographic evidence of pulmonary infiltrates compatible with pneumonia and at least one acute lower respiratory tract infection symptom or sign: cough, chest pain, dyspnea, fever >38° C, hypothermia <35°C, or abnormal breath sounds not explained by other causes;3. Hospitalization for ≥48 hours after admission through the emergency department due to CAP.Exclusion criteria included: 1. Pregnancy;2. Presence of respiratory failure prior to admission;3. Hospitalization primarily due to other more severe internal medical conditions not related to pneumonia;4. Death within 48 hours of admission;5. Presence of primary or metastatic malignancies involving the lungs or pleura;6. Concurrent malignancy undergoing active treatment.

### 2.2 Data Collection

Clinical data were collected from the hospital’s electronic medical records system, including: (1)Baseline characteristics:age, sex; (2) Vital signs at admission:heart rate, respiratory rate, temperature, systolic and diastolic blood pressure, and mental status; (3) Comorbidities:chronic pulmonary diseases, hypertension, coronary artery disease, cerebrovascular disease, congestive heart failure, renal dysfunction, myocardial infarction, diabetes, and history of malignancy (excluding respiratory system malignancies); (4) Laboratory tests:levels of transaminases, lactate dehydrogenase, triglycerides, total bilirubin, direct and indirect bilirubin, albumin, globulin, creatinine, blood urea nitrogen, serum sodium, potassium, calcium, phosphorus, lactate, serum glucose, N-terminal pro-brain natriuretic peptide (NT-proBNP), D-dimer, total white blood cell count, neutrophil percentage, red blood cell count, red cell distribution width, hemoglobin, platelet count, and albumin-to-globulin ratio; (5) Inflammatory and cytokine markers:C-reactive protein (CRP), procalcitonin (PCT); cytokines included tumor necrosis factor-α (TNF-α), interleukin-1β (IL-1β), IL-6, IL-8, and IL-10; (6) Radiographic findings: presence of pleural effusion on chest imaging.

### 2.3 Outcome Definition

The primary outcome was the development of respiratory failure within 48 hours of hospital admission. Respiratory failure was defined by meeting at least one of the following criteria:1. Documentation of respiratory failure (including “respiratory failure” or “acute respiratory failure”) in the clinical records; 2. Arterial blood gas analysis showing PaO₂ ≤ 60 mmHg or an acute increase in PaCO₂ ≥ 45 mmHg within 48 hours of admission; 3. Clinical records indicating the use of high-flow oxygen therapy or mechanical ventilation.

### 2.4 Data Preprocessing

Samples with >20% missing data were excluded. For variables with missing values, multiple imputation was performed using the ‘mice’ package (version 3.17.0) in R. Outliers in clinical and laboratory data—such as vital signs and biochemical values—were retained, as they may hold significant clinical meaning. Some lab test results that exceeded detectable limits were recorded in the electronic system using symbols such as “>” or “<” (e.g., PCT > 50pg/mL). Since such symbolic values cannot be processed by machine learning algorithms, we replaced these values with the respective maximum or minimum detectable limit values.

### 2.5 Model Development and Validation

We employed several supervised learning algorithms to construct predictive models: multivariable logistic regression (MLR), k-nearest neighbors (KNN), support vector machine (SVM), extreme gradient boosting (XGBoost), random forest (RF), and Light Gradient Boosting Machine (LightGBM). Patients were randomly divided into a training set and a test set at a 7:3 ratio. Ten-fold cross-validation was used during model training to prevent overfitting and enhance generalizability. Model performance was assessed using the area under the receiver operating characteristic curve (AUROC), and clinical utility was evaluated through decision curve analysis (DCA).Additionally, we developed an ensemble model by averaging the three top-performing models on the test set using a weighted approach. This ensemble strategy was intended to reduce bias introduced by the inherent limitations of any single algorithm and improve prediction performance for respiratory failure.

## 3. Statistical Analysis

All data processing and statistical analyses were performed using R software (version 4.1.3). Categorical variables were expressed as counts and percentages (N, %) and compared between groups using the chi-square test. For continuous variables, the distribution was first assessed for normality. Normally distributed variables were presented as mean (standard deviation (SD))and compared using the t-test. Non-normally distributed variables were reported as median and interquartile range (IQR: Q1-Q3) and analyzed using the Mann–Whitney U test. A p-value < 0.05 was considered statistically significant.

## 4. Results

### 4.1 Baseline Characteristics

A total of 257 patients met the inclusion and exclusion criteria and were included in the final analysis for predicting the risk of acute respiratory failure (ARF) within 48 hours of admission for community-acquired pneumonia (CAP). The median age was 76.0 years (IQR: 68.0–84.0), and 145 (56.4%) were male. Among them, 148 patients (57.6%) developed ARF within 48 hours of admission. Baseline characteristics are summarized in Table 1.

**Table 1.** Baseline characteristics.

| Variables | All(N=257) | Negative Group<br>(N=109) | Positive Group<br>(N=148) | P_value |
| --- | --- | --- | --- | --- |
| Demographic data |  |  |  |  |
| Gender = Male (%) | 145 (56.4) | 61 (56.0) | 84 (56.8) | 1.000 |
| Age (year) | 76.00 [68.00, 84.00] | 78.00 [69.00, 85.00] | 75.00 [67.75, 84.00] | 0.313 |
| Vital Signs |  |  |  |  |
| Heart_rate (beats/min) | 98.00 [86.00, 112.00] | 91.00 [81.00, 103.00] | 103.50 [89.00, 114.25] | <0.001 |
| Respiratory_rate (breaths/min) | 20.00 [18.00, 22.00] | 18.00 [17.00, 20.00] | 21.00 [19.00, 23.00] | <0.001 |
| T (°C) | 36.60 [36.40, 37.00] | 36.60 [36.40, 37.30] | 36.60 [36.38, 36.80] | 0.257 |
| SBP (mmHg) | 136.63 (26.78) | 134.96 (27.47) | 137.86 (26.29) | 0.393 |
| DBP (mmHg) | 79.00 [67.00, 92.00] | 76.00 [66.00, 89.00] | 80.00 [68.75, 94.00] | 0.149 |
| Abnormal Consciousness = YES (%) | 48 (18.7) | 16 (14.7) | 32 (21.6) | 0.211 |
| Laboratory Results |  |  |  |  |
| Lac (mmol/L) | 1.30 [1.00, 1.80] | 1.20 [0.90, 1.80] | 1.40 [1.00, 1.83] | 0.143 |
| AST (U/L) | 28.00 [20.00, 41.00] | 28.00 [20.00, 39.00] | 28.00 [20.00, 45.75] | 0.574 |
| ALT (U/L) | 21.00 [14.00, 36.00] | 19.00 [13.00, 36.00] | 22.00 [14.00, 37.00] | 0.358 |
| LDH (U/L) | 245.00 [204.00, 349.00] | 232.00 [193.00, 318.00] | 268.00 [211.25, 356.25] | 0.025 |
| TG (mmol/L) | 1.07 [0.79, 1.46] | 1.07 [0.79, 1.45] | 1.08 [0.80, 1.48] | 0.827 |
| TBIL (umol/L) | 11.40 [7.90, 17.70] | 11.00 [8.00, 13.80] | 12.40 [7.90, 21.42] | 0.062 |
| DBIL (umol/L) | 5.00 [3.50, 8.00] | 4.70 [3.70, 6.30] | 5.10 [3.38, 9.22] | 0.208 |
| Alb (g/L) | 35.88 (5.63) | 36.10 (5.75) | 35.71 (5.55) | 0.588 |
| Glb (g/L) | 27.60 [24.00, 32.00] | 28.00 [25.00, 32.00] | 26.85 [23.78, 31.00] | 0.050 |
| Creatine<br>(umol/L) | 76.80 [59.70, 115.20] | 82.10 [56.00, 122.00] | 76.10 [61.40, 113.88] | 0.786 |
| BUN<br>(mmol/L) | 8.20 [6.21, 12.44] | 8.03 [6.14, 12.16] | 8.27 [6.28, 12.47] | 0.557 |
| Na (mmol/L) | 137.60 [133.80, 140.20] | 137.80 [134.60,<br>140.20] | 137.50 [133.33, 139.93] | 0.587 |
| K (mmol/L) | 4.10 [3.80, 4.50] | 4.10 [3.80, 4.40] | 4.10 [3.70, 4.50] | 0.920 |
| Ca (mmol/L) | 2.07 [1.96, 2.17] | 2.05 [1.94, 2.17] | 2.08 [2.00, 2.18] | 0.103 |
| P (mmol/L) | 1.03 [0.84, 1.31] | 1.02 [0.85, 1.32] | 1.04 [0.84, 1.27] | 0.845 |
| Glu (mmol/L) | 7.84 [6.16, 10.46] | 7.58 [6.12, 9.44] | 8.50 [6.21, 11.10] | 0.134 |
| Nt_BNP<br>(pg/mL) | 1904.00 [309.00,<br>4918.00] | 1795.00 [201.00,<br>4818.00] | 1927.00 [466.75,<br>4947.50] | 0.467 |
| D_dimer<br>(median<br>[IQR]) | 1.18 [0.43, 2.93] | 0.90 [0.39, 2.39] | 1.36 [0.50, 3.52] | 0.036 |
| PT (s) | 13.20 [12.40, 14.60] | 13.30 [12.30, 14.80] | 13.20 [12.50, 14.40] | 0.589 |
| APTT (s) | 29.60 [26.70, 33.10] | 29.70 [26.80, 34.40] | 29.30 [26.58, 32.32] | 0.245 |
| INR | 1.08 [1.00, 1.22] | 1.08 [0.98, 1.22] | 1.08 [1.01, 1.21] | 0.682 |
| WBC (K/mcL) | 9.43 [6.98, 13.82] | 8.72 [6.85, 14.46] | 10.00 [7.27, 13.81] | 0.508 |
| NEUT% | 84.40 [76.30, 90.20] | 83.90 [75.60, 90.00] | 84.60 [77.20, 90.62] | 0.602 |
| RBC (M/mcL) | 3.96 [3.43, 4.46] | 4.05 [3.63, 4.51] | 3.92 [3.33, 4.40] | 0.182 |
| HCT (%) | 35.60 [30.60, 40.50] | 36.50 [32.40, 41.10] | 35.25 [29.78, 39.85] | 0.158 |
| Hb (g/L) | 118.52 (27.53) | 120.13 (27.73) | 117.34 (27.40) | 0.423 |
| Plt (K/mcL) | 195.00 [143.00, 266.00] | 201.00 [157.00,<br>283.00] | 188.50 [138.25, 252.25] | 0.086 |
| AG_Ratio | 1.32 (0.32) | 1.30 (0.32) | 1.34 (0.31) | 0.227 |
| Inflammatory Markers |  |  |  |  |
| CRP (mg/L) | 47.00 [13.00, 99.00] | 40.00 [14.00, 106.00] | 50.00 [12.50, 96.25] | 0.904 |
| PCT (ng/mL) | 0.07 [0.05, 0.97] | 0.06 [0.05, 0.86] | 0.14 [0.05, 1.05] | 0.202 |
| IL1_beta<br>(pg/mL) | 4.54 [2.14, 7.10] | 2.44 [1.01, 6.16] | 5.67 [2.47, 7.35] | <0.001 |
| IL_6 (pg/mL) | 22.37 [8.33, 64.66] | 19.11 [7.34, 47.11] | 24.63 [8.94, 77.49] | 0.256 |
| IL_8 (pg/mL) | 14.97 [7.53, 33.53] | 14.97 [4.53, 35.89] | 15.00 [8.71, 30.56] | 0.419 |
| IL_10 (pg/mL) | 2.03 [1.29, 3.77] | 2.27 [1.40, 3.83] | 1.85 [1.19, 3.58] | 0.169 |
| TNF_aphla<br>(pg/mL) | 2.35 [0.65, 4.86] | 1.16 [0.52, 2.99] | 3.40 [0.93, 5.25] | <0.001 |
| GCS (%) |  |  |  | 0.205 |
| Mild_coma | 202 (78.6) | 91 (83.5) | 111 (75.0) |  |
| Moderate_co<br>ma | 46 (17.9) | 16 (14.7) | 30 (20.3) |  |
| Severe_coma | 9 (3.5) | 2 (1.8) | 7 (4.7) |  |
| Pleural_Effusi<br>on = YES (%) | 94 (36.6) | 28 (25.7) | 66 (44.6) | 0.003 |
| Vasoactive_D<br>rug = YES<br>(%) | 15 (5.8) | 5 (4.6) | 10 (6.8) | 0.643 |
| Comorbidities |  |  |  |  |
| Chronic_lung<br>_disease =<br>YES (%) | 59 (23.0) | 30 (27.5) | 29 (19.6) | 0.179 |
| Hypertension<br>= YES (%) | 161 (62.6) | 69 (63.3) | 92 (62.2) | 0.955 |
| Coronary_he<br>art_disease =<br>YES (%) | 90 (35.0) | 40 (36.7) | 50 (33.8) | 0.725 |
| Heart_Disfunction = YES (%) | 65 (25.3) | 25 (22.9) | 40 (27.0) | 0.548 |
| Renal_Disfunction = YES (%) | 45 (17.5) | 18 (16.5) | 27 (18.2) | 0.846 |
| Heart_attack = YES (%) | 13 (5.1) | 5 (4.6) | 8 (5.4) | 0.994 |
| Cerebrovascular_disease = YES (%) | 44 (17.1) | 16 (14.7) | 28 (18.9) | 0.469 |
| Diabetes = YES (%) | 96 (37.4) | 34 (31.2) | 62 (41.9) | 0.105 |
| Malignant_tumour = YES (%) | 35 (13.6) | 11 (10.1) | 24 (16.2) | 0.218 |

Data are presented as Mean (SD) for normally distributed continuous variables, Median (IQR) for non-normally distributed continuous variables, and counts (percentages) for categorical variables in the Total, Negative group, and Positive group columns. SD, standard deviation; IQR, interquartile range.
Abbreviations: T, temperature; SBP, systolic blood pressure; DBP, diastolic blood pressure; Lac, lactate; AST, aspartate aminotransferase; ALT, alanine aminotransferase; LDH, lactate dehydrogenase; TG, triglycerides; TBIL, total bilirubin; DBIL, direct bilirubin; Alb, albumin; Glb, globulin; BUN, blood urea nitrogen; Na, serum sodium; K, serum potassium; Ca, serum calcium; P, serum phosphorus; Glu, glucose; Nt\_BNP, N-terminal pro-brain natriuretic peptide; PT, prothrombin time; APTT, activated partial thromboplastin time; INR, international normalized ratio; WBC, white blood cell count; NEUT%, neutrophil percentage; RBC, red blood cell count; HCT, hematocrit; Hb, hemoglobin; Plt, platelet count; AG\_Ratio, albumin-to-globulin ratio; CRP, C-reactive protein; PCT, procalcitonin; IL, interleukin; TNF, tumor necrosis factor; GCS, Glasgow Coma Scale.

Regarding vital signs, patients in the ARF group (positive group) had a significantly higher respiratory rate than those in the non-ARF group (negative group) (21.0 vs. 18.0 breaths/min, p < 0.001), and heart rate was also significantly higher in the ARF group (103.5 vs. 91.0 beats/min, p < 0.001). In laboratory parameters, the ARF group had significantly higher levels of lactate dehydrogenase (268 vs. 232 U/L, p = 0.025), lower levels of immunoglobulin (26.85 vs. 28.00 g/L, p = 0.050), and elevated D-dimer (1.36 vs. 0.90 mg/L, p = 0.036). Inflammatory markers including IL-1β (5.67 vs. 2.44 pg/mL, p < 0.001) and TNF-α (3.40 vs. 1.16 pg/mL, p < 0.001) were significantly higher in the ARF group. No significant differences were observed between the two groups regarding baseline comorbidities.

### 4.2 Model Development

To identify key predictive features associated with acute respiratory failure (ARF) within 48 hours of admission, Least Absolute Shrinkage and Selection Operator (LASSO) regression was applied to all candidate variables (Figure 1A and 1B). LASSO addresses multicollinearity and prevents overfitting by penalizing less informative variables, effectively shrinking their coefficients toward zero. At the optimal regularization parameter (λ = 0.045), LASSO selected eight features: respiratory rate, tumor necrosis factor-α (TNF-α), pleural effusion, temperature, interleukin-1 β (IL-1β), heart rate, total bilirubin, and interleukin-8 (IL-8). These features were then entered into a multivariable logistic regression (MLR) model to adjust for potential confounders. The final MLR model retained six predictors: respiratory rate, TNF-α, pleural effusion, temperature, IL-1β, and heart rate.

**Figure 1A.**
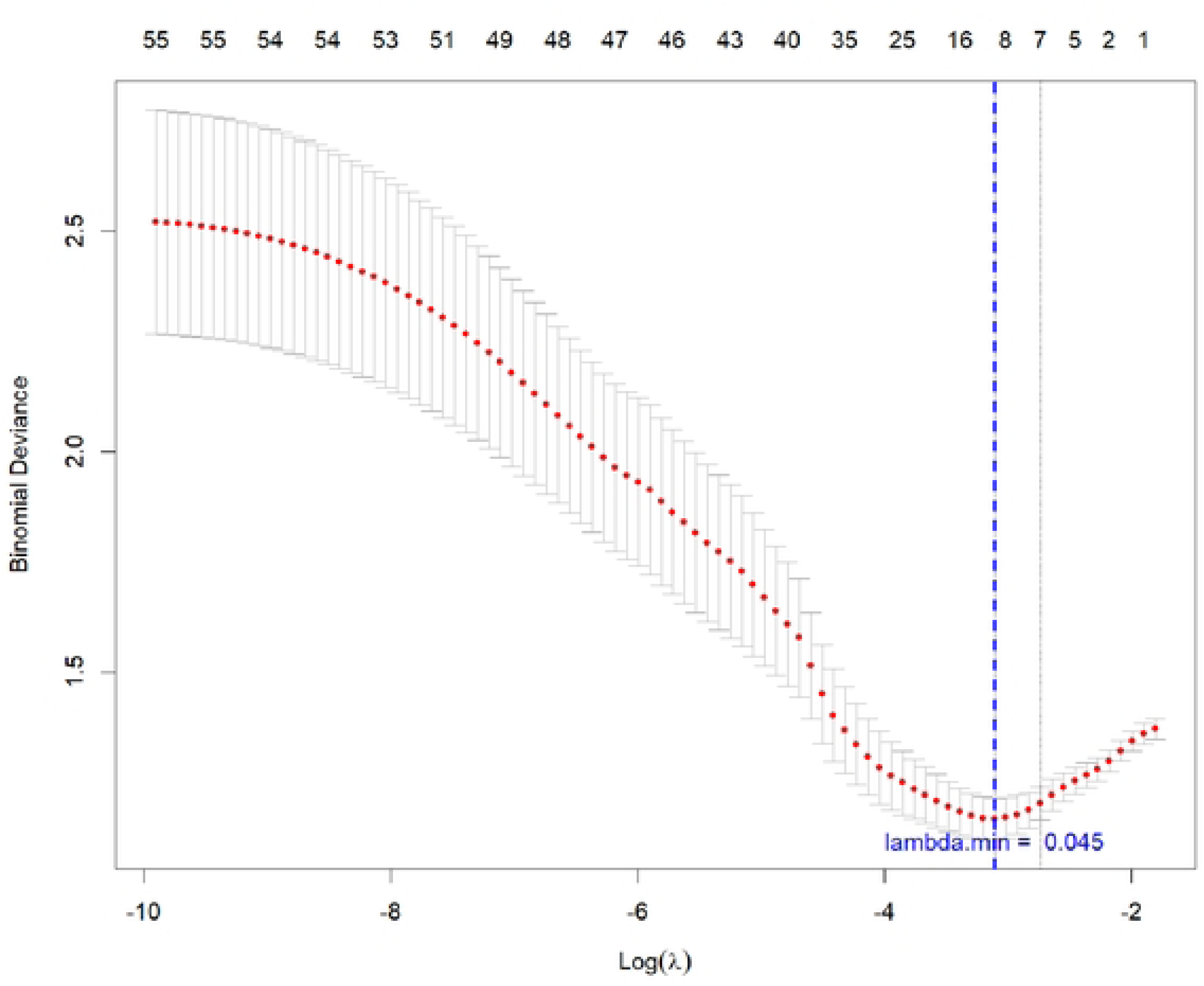
illustrates the variation in binomial deviance across different levels of regularization strength (Log(λ)) during lasso regression model training using 10-fold cross-validation. The red dots represent the mean deviance at each λ value, while the grey error bars indicate the corresponding standard errors. The blue dashed line marks the X value that minimizes the mean deviance {lambda.min = 0.045), indicating the model with the optimal performance on the validation set.

**Figure 1B.**
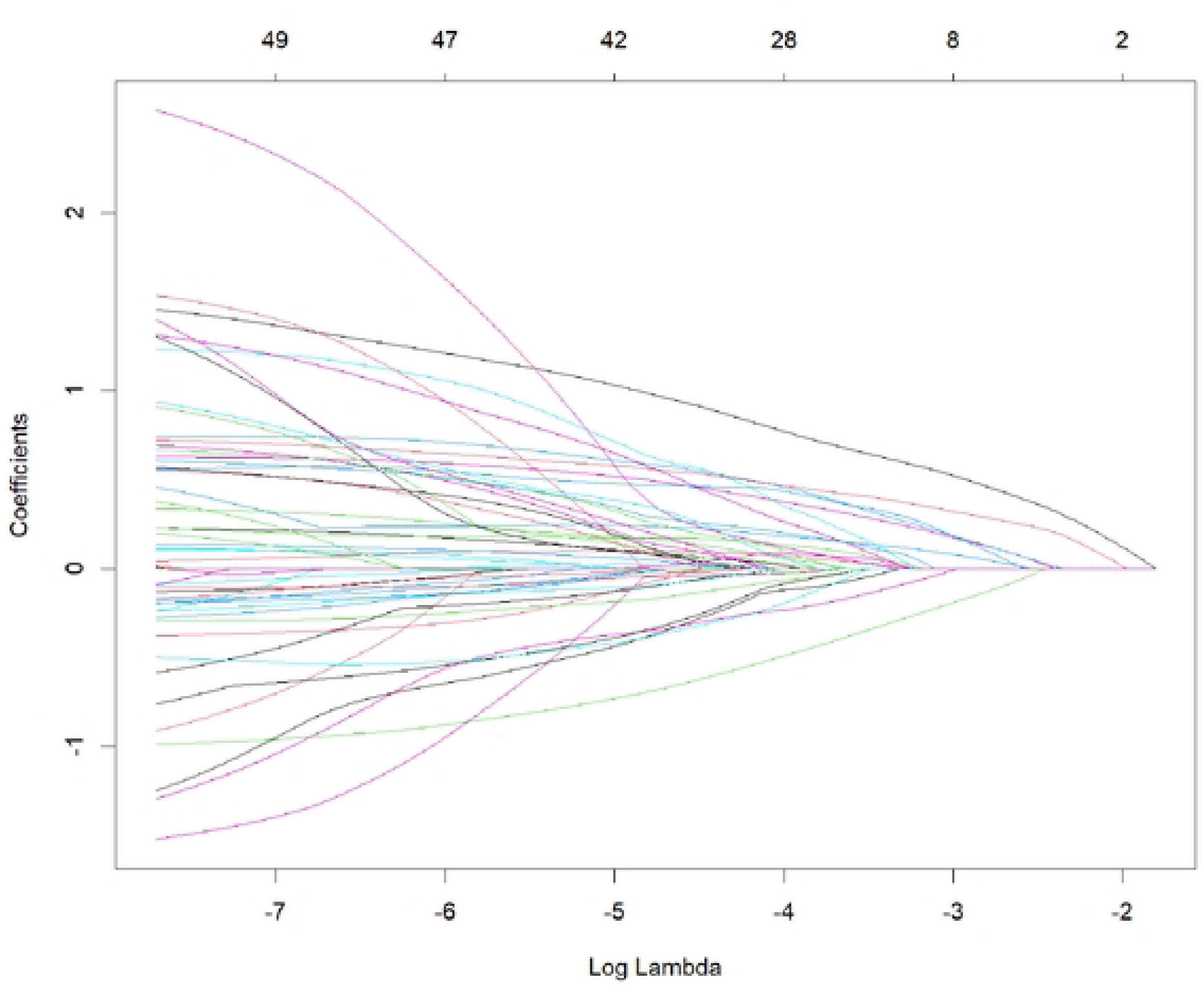
Coefficient paths in Lasso regression under varying values of the regularization parameter λ, with the x-axis representing the logarithm of λ (Log Lambda). Each curve corresponds to the trajectory of a predictor’s coefficient as λ changes. As λ increases (moving leftward), more coefficients aregradually shrunk toward zero.

To leverage the ability of machine learning (ML) algorithms to capture complex nonlinear interactions among variables, we trained five ML models using the full feature set: k-nearest neighbors (KNN), support vector machine (SVM), extreme gradient boosting (XGBoost), Light Gradient Boosting Machine (LightGBM), and random forest (RF). Model performance in both training and test datasets is presented in Figure 2A and 2B.

**Figure 2A.**
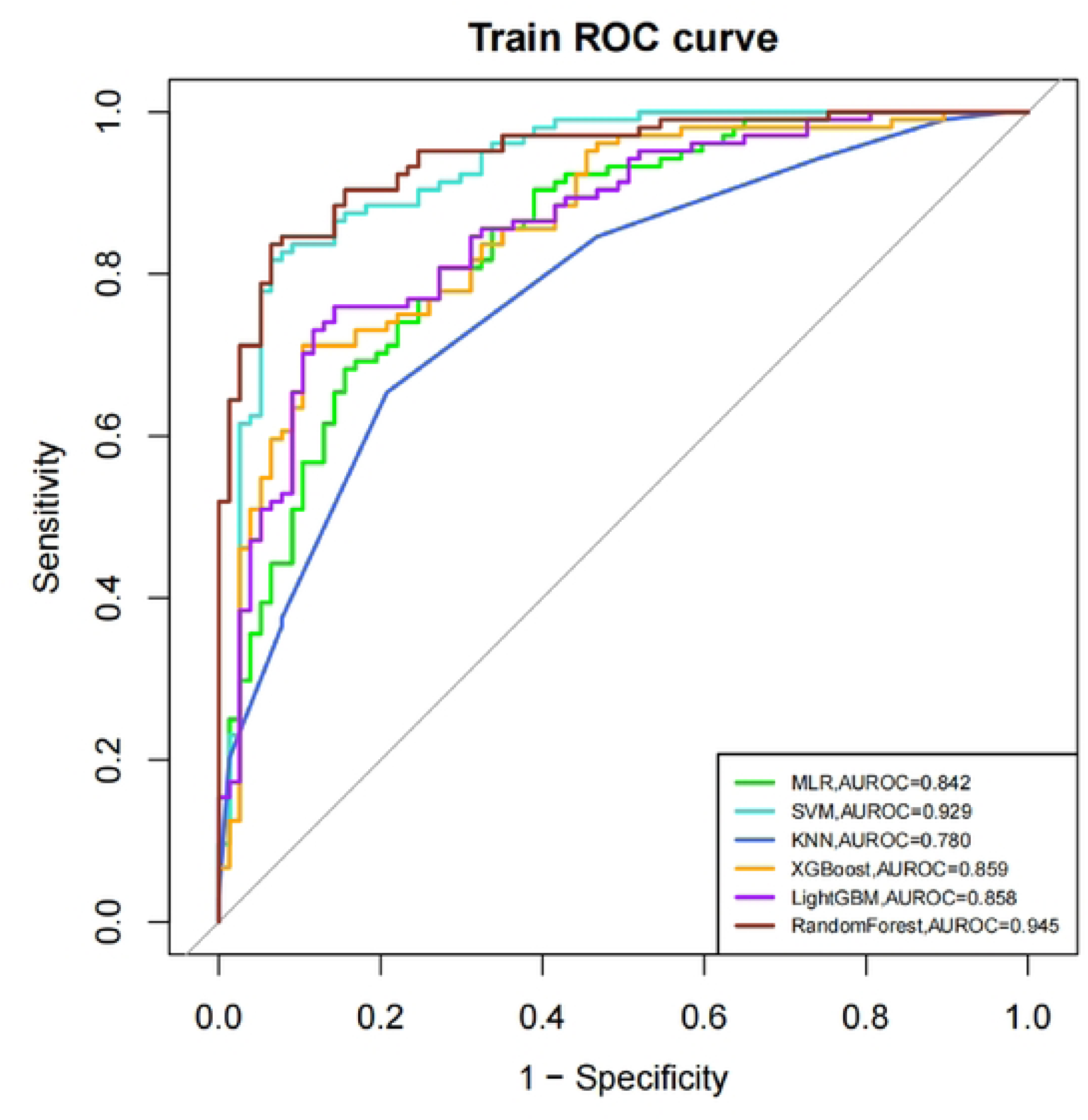
ROC Curves for Different Models on the Training Set. This figure shows the Receiver Operating Characteristic (ROC) curves for six classification models evaluated on the training set. The x-axis represents 1 – specificity (false positive rate), and the y-axis represents sensitivity (true positive rate). Each colored curve corresponds to a specific model, with the Area Under the ROC Curve (AUROC) values listed in the legend. The models with the highest performance were Random Forest (AUROC = 0.945), SVM (AUROC = 0.929), and XGBoost (AUROC = 0.858).

**Figure 2B.**
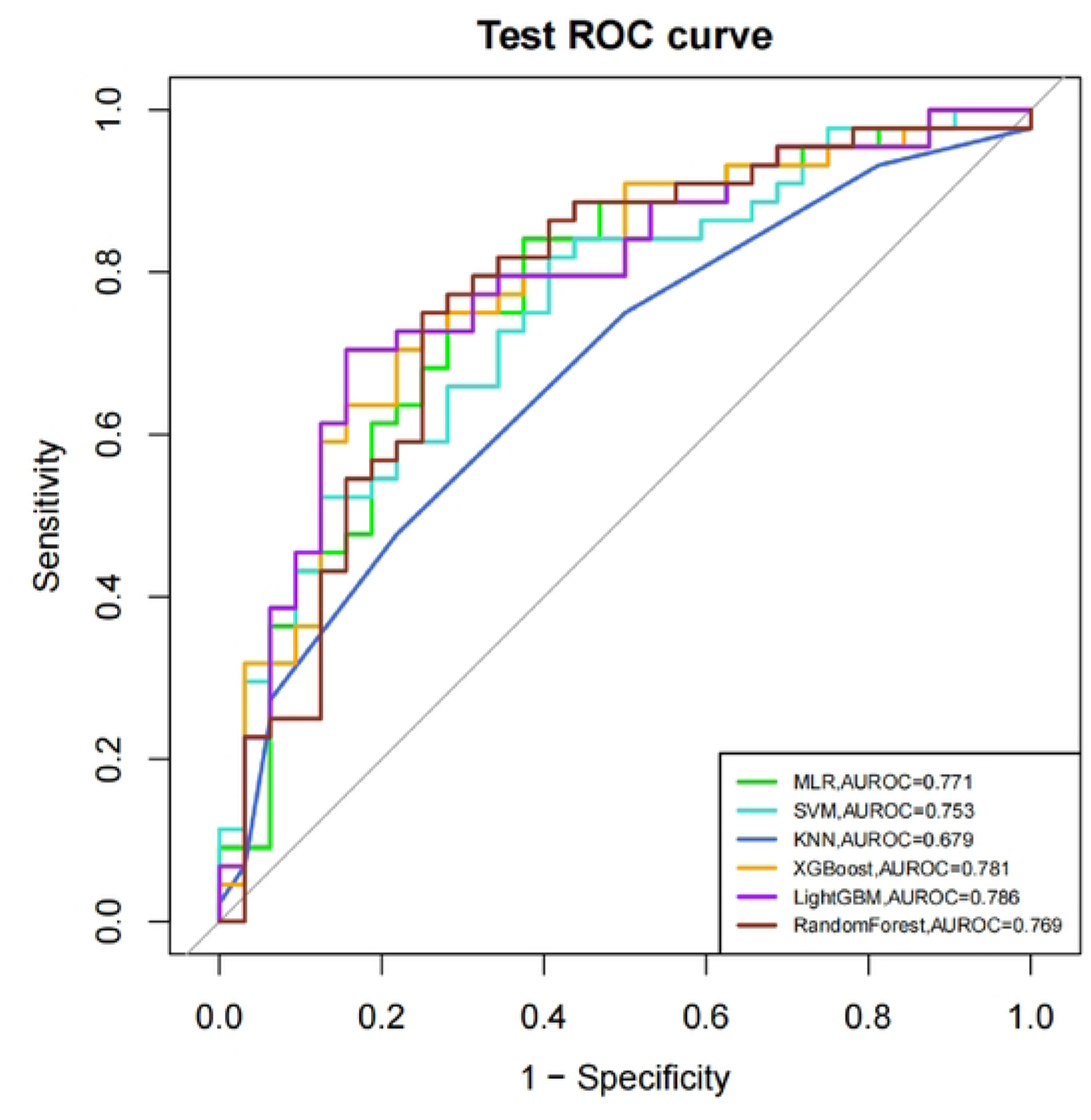
displays the ROC curves of each model evaluated on the test set. Compared to the training results, all models showed a decrease in AUROC values on the test set, which is a common occurrence when assessing model generalization. Among them, LightGBM (AUROC = 0. 786), XGBoost (AUROC = 0. 781), and multiple logistic regression (MLR, AUROC = 0. 771) achieved the best performance. The reductions in AUROC for these models were all less than 10%compared to their corresponding training set values, indicating good model fitting.

In the training set, the top-performing models were Random Forest (AUROC = 0.945), followed by SVM (AUROC = 0.929), and XGBoost (AUROC = 0.858). In the test set, performance declines were less than 10% for all models, suggesting good generalizability. The best-performing models in the test set were LightGBM (AUROC = 0.786), XGBoost (AUROC = 0.781), and MLR (AUROC = 0.771).

To evaluate the clinical utility of these models across different risk thresholds, decision curve analysis (DCA) was performed on the top three models in the test set (Figure 3A). Across most threshold ranges, all three models demonstrated superior net benefit compared to the “treat all” and “treat none” strategies, with comparable performance among the models, thereby supporting their utility in risk-based clinical decision-making.

**Figure 3A.**
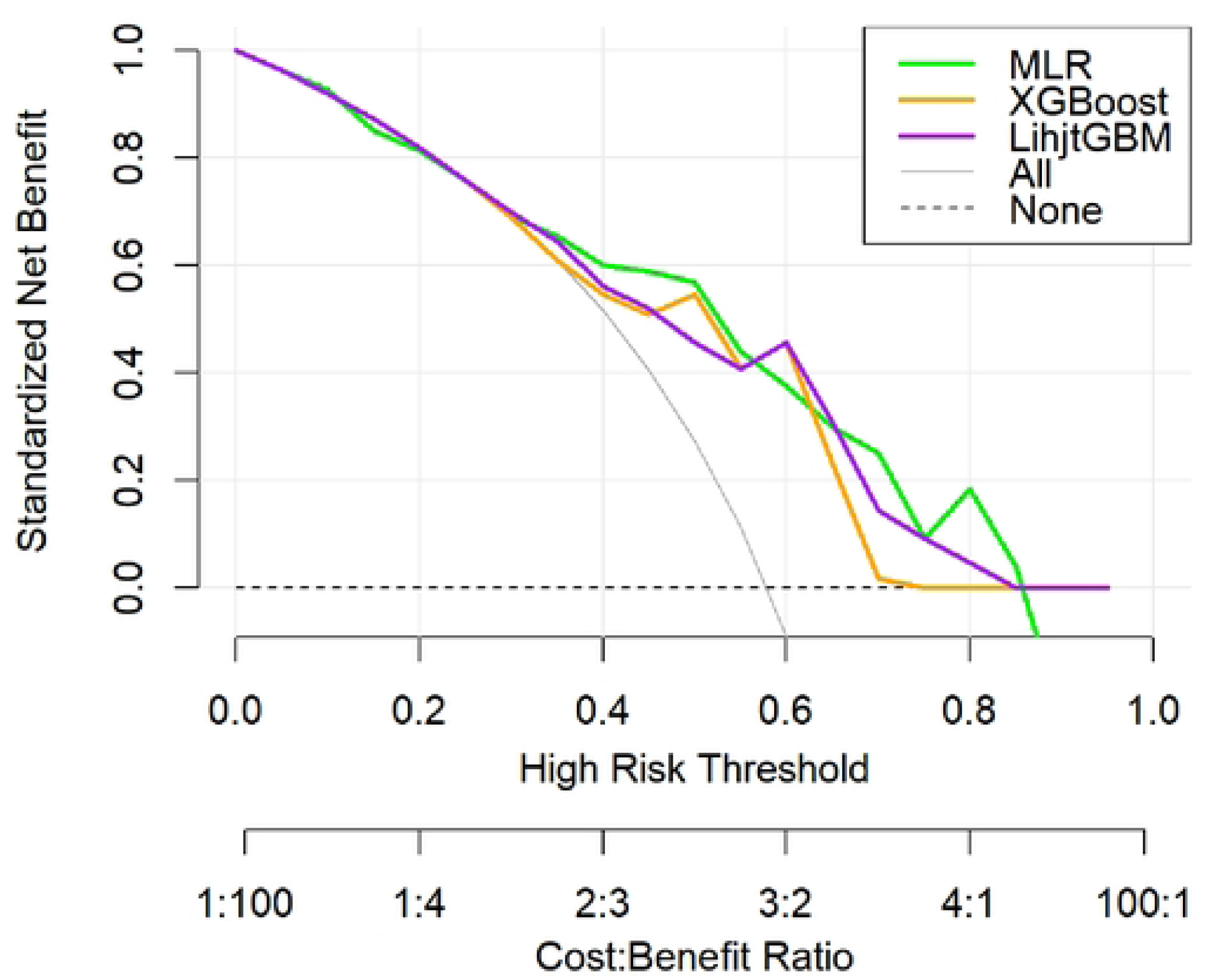
This figure illustrates the standardized net benefit of multiple logistic regression (MLR), XGBoost, and LightGBM across a range of high-risk threshold probabilities. The x-axis represents the threshold for classifying individuals as high risk, with the corresponding cost-benefit ratios shown below. The y-axis shows the standardized net benefit. The “All” curve assumes all individuals are treated as high risk, while the “None” line indicates no individuals are treated. Across most ranges of risk thresholds, the net benefits of all three models exceed those of the “All” and “None” strategies, andtheir net benefit curves are generally similar.

**Figure 3B.**
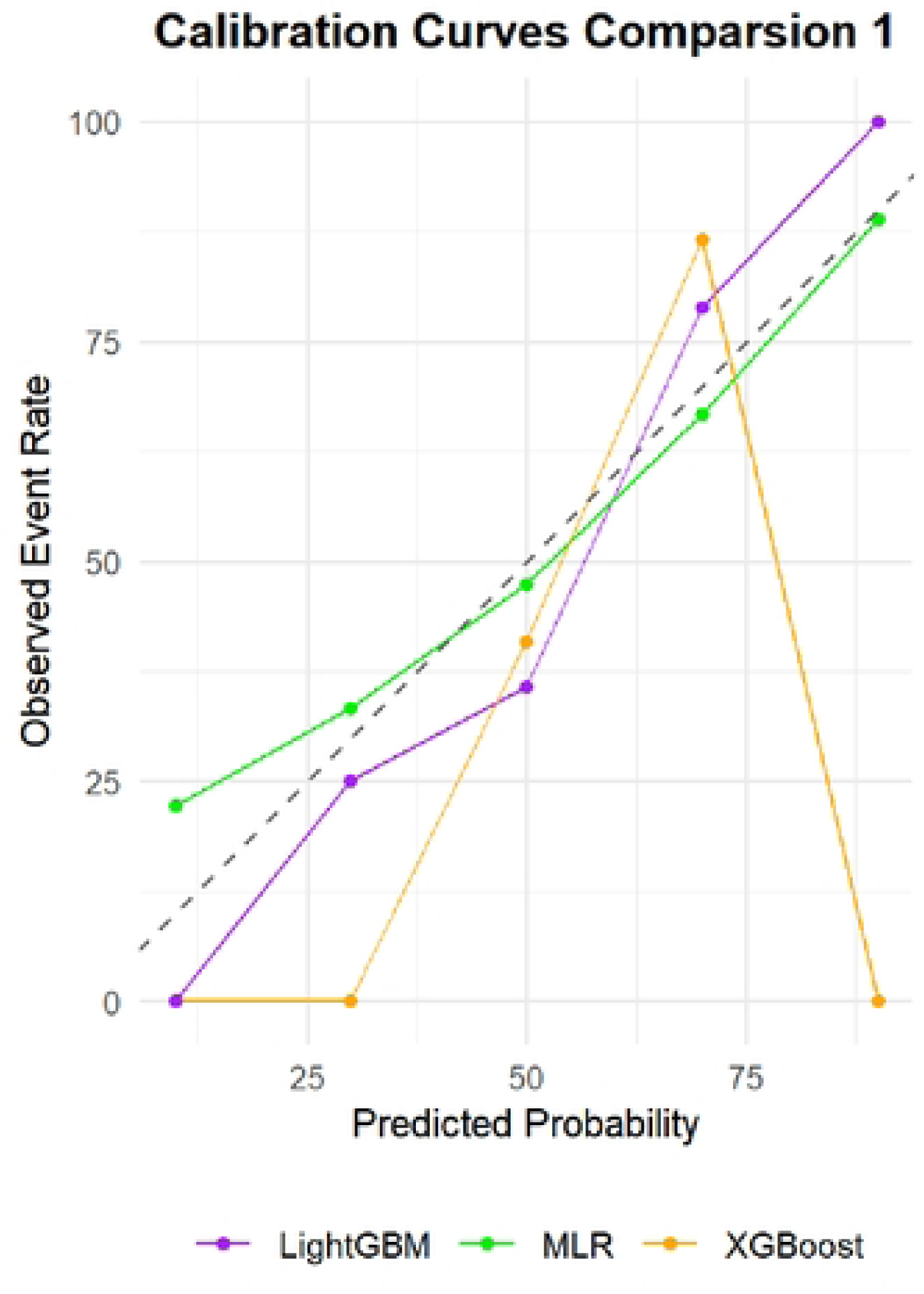
The plot shows the calibration performance of the LightGBM, XGBoost, and multiple logistic regression (MLR) models, comparing predicted probabilities with observed event rates. The x-axis represents the predicted probability of an event, while the y-axis indicates the actual observed event rate. The dashed diagonal line represents perfect calibration, where predicted probabilities exactly match observed outcomes. The MLR and LightGBM models demonstrate better agreement between predicted and observed probabilities on the test set, indicating superior calibration. In contrast, the XGBoost model shows noticeable deviation in the high-probability range, suggesting potential overestimationin those regions.

**Figure 3C.**
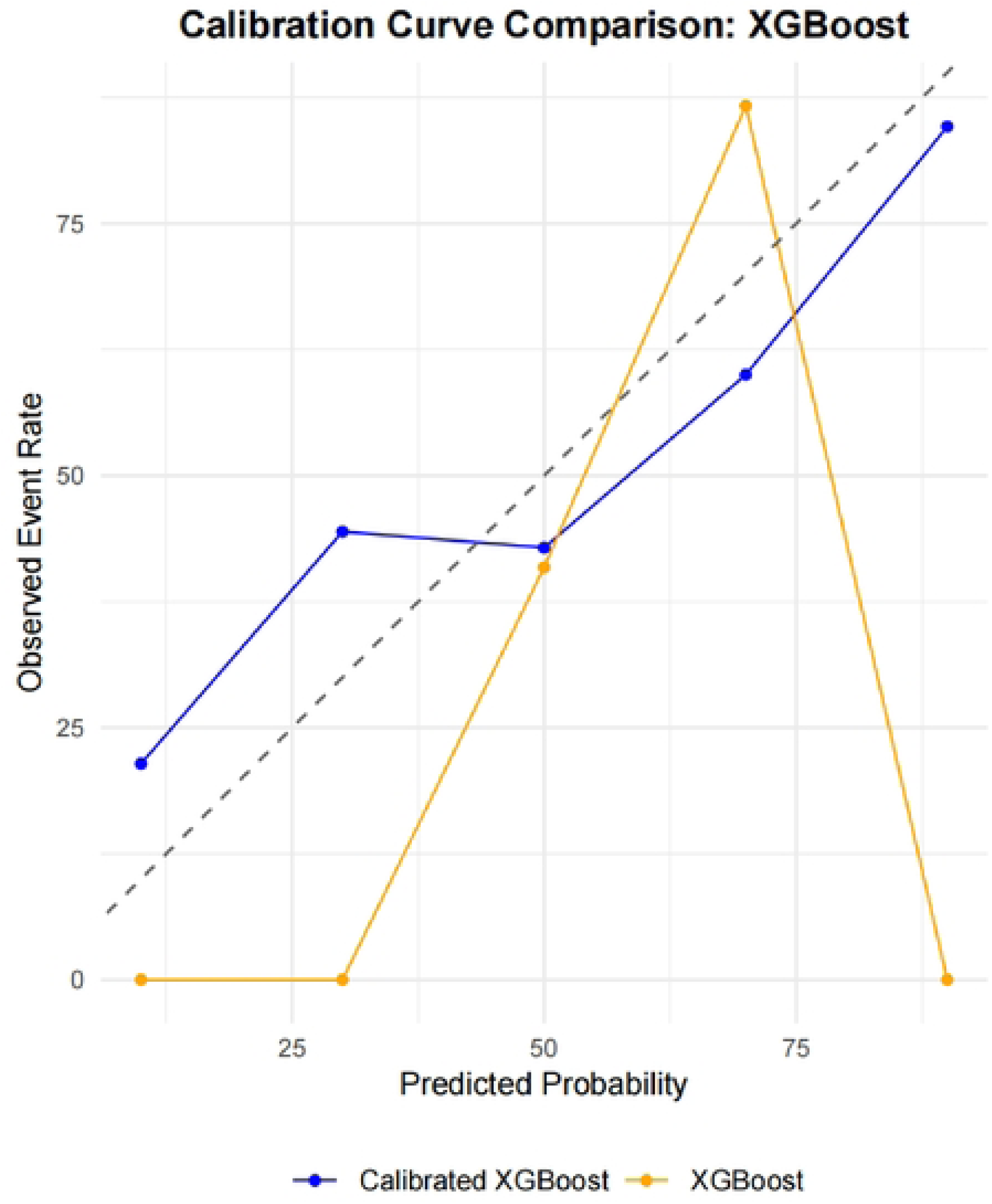
Calibration performance of the original XGBoost model (orange) and the Platt-scaled XGBoost model (blue) across different predicted probability intervals. After Platt scaling, the calibration of XGBoostl improved notably across all probability ranges, with the blue curve aligning more closely with the ideal calibration line.

Calibration performance was assessed by plotting calibration curves for the LightGBM, MLR, and XGBoost models (Figure 3B). The MLR and LightGBM models exhibited relatively smooth and well-aligned calibration curves. In contrast, the XGBoost model showed greater variability and substantial deviations, particularly in the high predicted probability range.

To correct calibration bias in the XGBoost model, post hoc calibration was performed using Platt scaling—a logistic regression-based method. Because tree-based ensemble models such as XGBoost are prone to overfitting, especially in small datasets, Platt scaling helps by fitting a logistic function to the predicted scores. The recalibrated XGBoost model demonstrated a notably improved calibration curve, particularly in high-risk predictions, aligning more closely with the ideal reference line (Figure 3C), thereby reducing overprediction.

Based on these findings, LightGBM, MLR, and the calibrated XGBoost model were selected for ensemble modeling using a weighted averaging strategy. This approach was motivated by several considerations:First, each model contributes distinct strengths. Tree-based models (LightGBM and XGBoost) excel at capturing nonlinear relationships, while MLR offers high interpretability and robust performance with linear associations.By combining models of different types, the ensemble captures a broader range of data patterns and compensates for the individual limitations of each method.Ensemble learning also enhances robustness against performance degradation caused by distributional shifts.Finally, given the relatively small sample size, a simple weighted averaging method was chosen to reduce computational complexity and mitigate overfitting, while maintaining strong predictive performance.

The weighted ensemble model was constructed by integrating predictions from LightGBM, MLR, and the recalibrated XGBoost model. Figure 4A presents the receiver operating characteristic (ROC) curves for the ensemble and its three constituent models. The ensemble achieved an AUROC of 0.792 (95% CI: 0.682– 0.897), with an accuracy of 0.75 (95% CI: 0.637–0.842), sensitivity of 0.813, specificity of 0.705, positive predictive value (PPV) of 0.667, and negative predictive value (NPV) of 0.838. Calibration performance of the ensemble and base models is shown in Figure 4B. A comparison of predictive performance metrics across all four models on the test set is provided in Table 2.

**Figure 4A.**
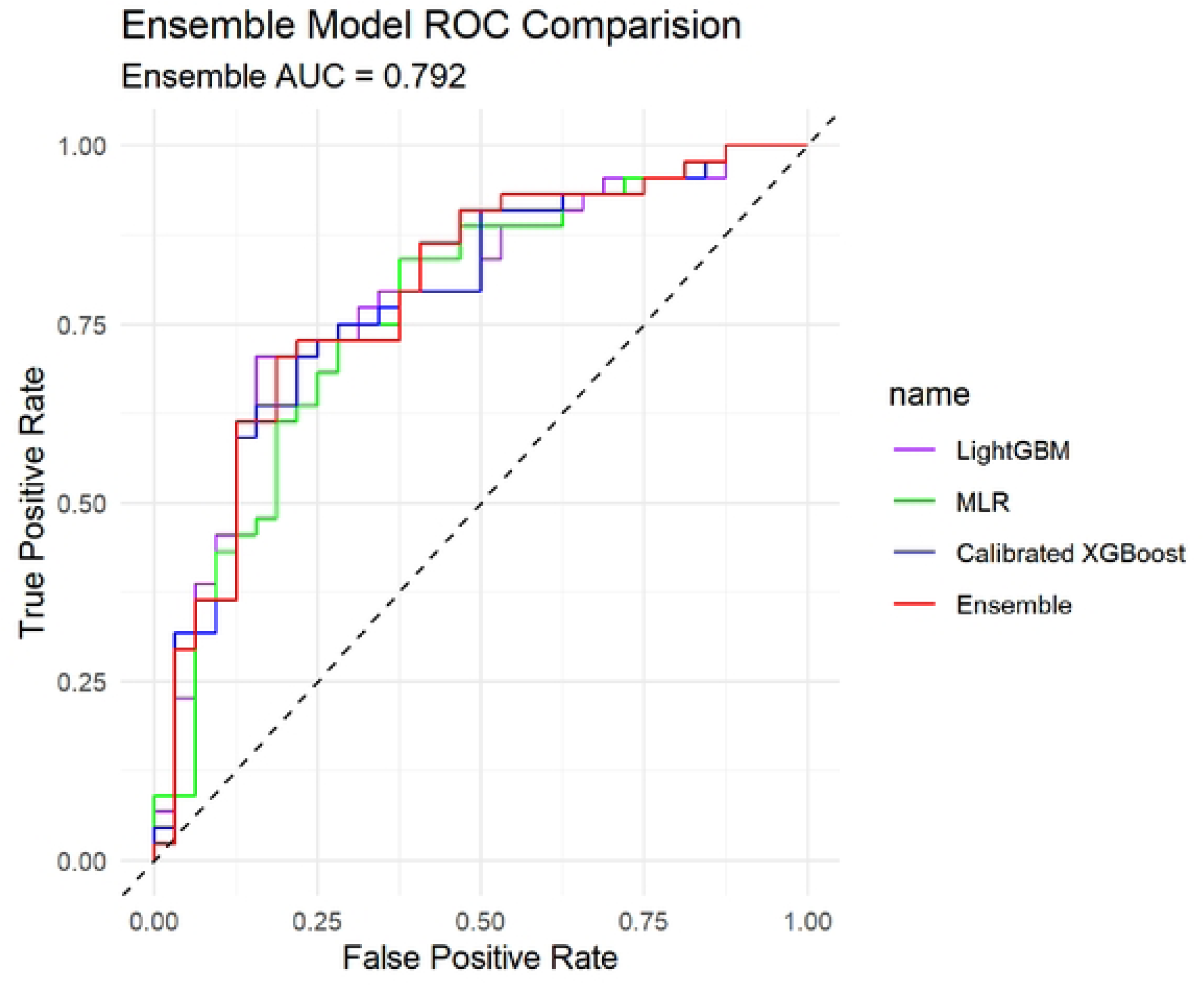
ROC curve comparison of the ensemble model and base models on the test set. The plot displays the receiver operating characteristic (ROC) curves for LightGBM (purple), multiple logistic regression (MLR, green), Platt-calibrated XGBoost (blue), and their ensemble model (red). The ensemble model (red curve) outperforms each individual model overall, with an AUC of 0.792.

**Figure 4B.**
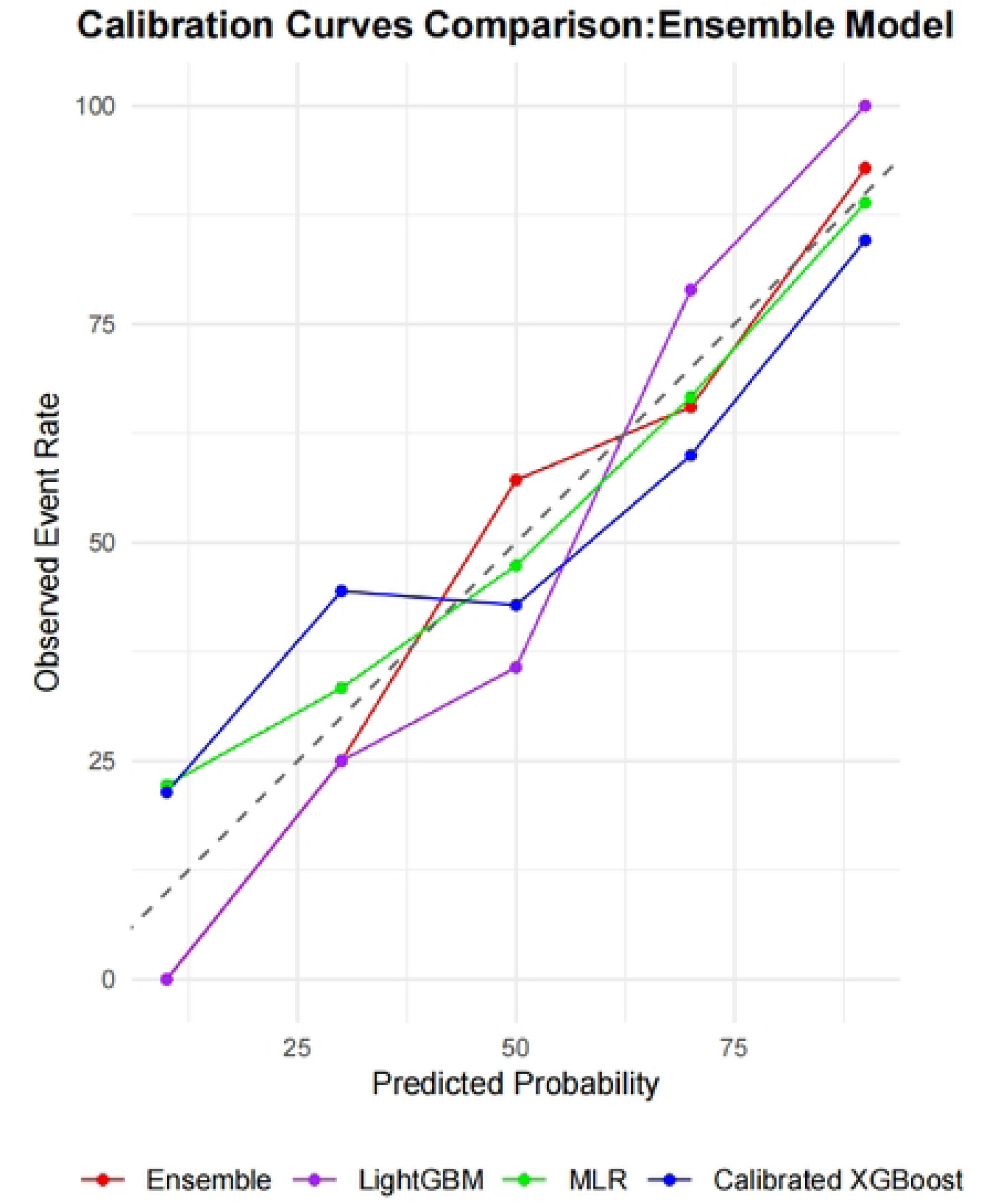
Calibration curve comparison of the ensemble model and base models on the test set. The figure shows the observed event rates across different predicted probability intervals for the ensemble model (red), LightGBM (purple), multiple logistic regression (MLR, green), and Platt-calibrated XGBoost (blue). The ensemble model demonstrates better or comparable calibration performance across multiple probability intervals compared to the individual models.

**Table 2.**
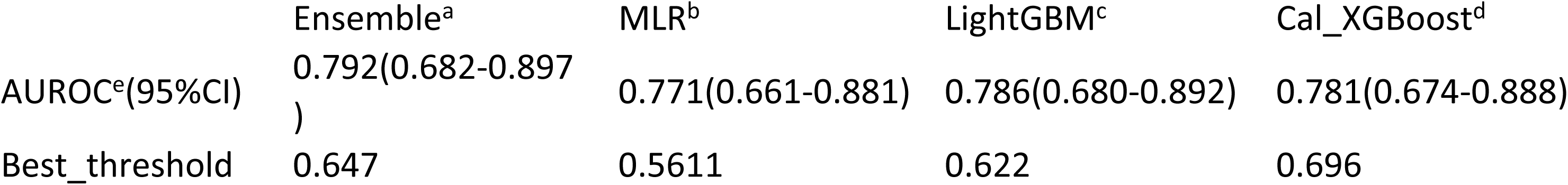
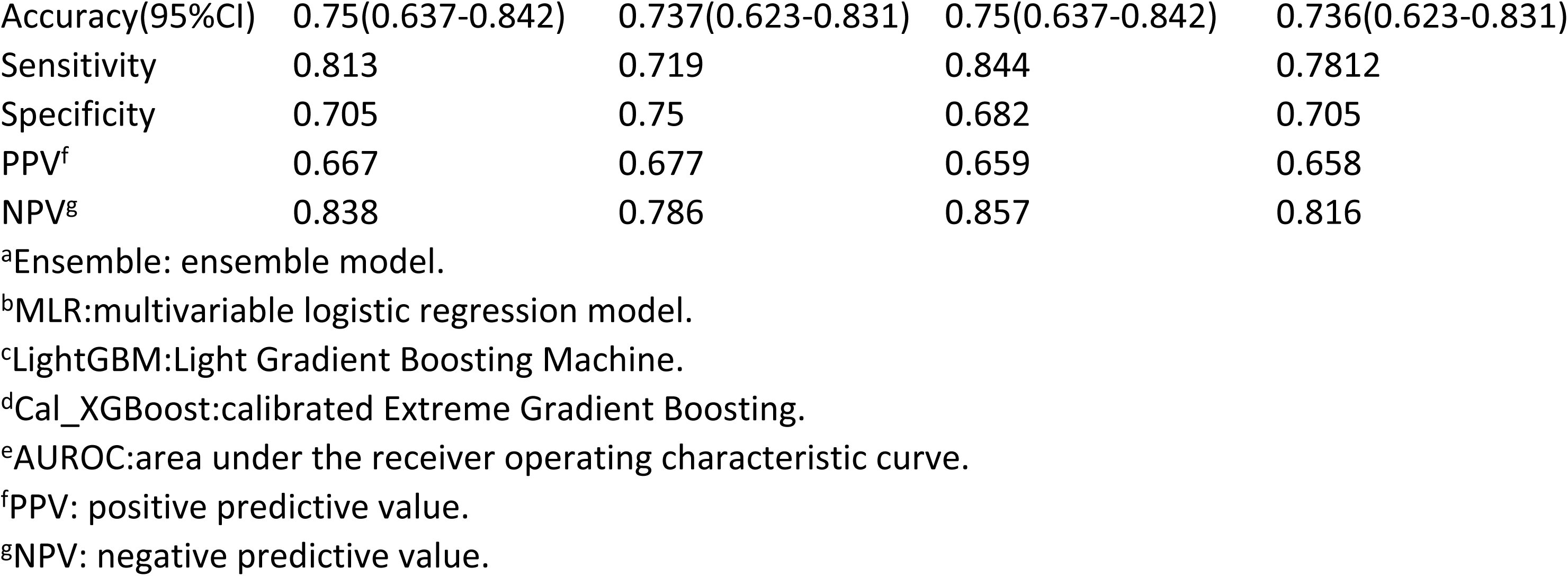
Performance of the ARF risk prediction models in the CAP testing Cohort.

### 4.3 Model Interpretability

To improve the interpretability of the ensemble model and identify key features associated with the onset of acute respiratory failure (ARF) within 48 hours of hospital admission in patients with community-acquired pneumonia (CAP), SHapley Additive exPlanations (SHAP) values were calculated. SHAP values offer both individual-level interpretability and global insights into feature importance and the directional impact of each variable on model predictions.

By ranking the mean absolute SHAP values, we identified the top ten most influential features contributing to the ensemble model’s predictions, as illustrated in Figure 5A. Respiratory rate was the most important predictor, followed by tumor necrosis factor-α (TNF-α), interleukin-1β (IL-1β), heart rate, and pleural effusion. Additional key features included body temperature, total bilirubin, serum calcium, albumin-to-globulin ratio, and platelet count.

A SHAP beeswarm plot (Figure 5B) visualizes the distribution of SHAP values for the top 10 features, showing their influence on individual predictions. Higher SHAP values reflect a stronger positive contribution to the predicted risk of ARF, while lower SHAP values indicate a negative association. An increased respiratory rate was consistently linked to higher SHAP values, significantly elevating the predicted ARF risk—highlighting its role as a crucial early warning sign. Similarly, elevated levels of TNF-α and IL-1β were strongly associated with increased risk, suggesting that heightened inflammatory activity may play a central role in the pathogenesis of early respiratory failure. Elevated heart rate, body temperature, and the presence of pleural effusion also contributed positively to the predicted likelihood of ARF.

## 5. Discussion

In this study, we developed and validated a multi-model, weighted ensemble prediction tool to assess the risk of acute respiratory failure (ARF) within 48 hours of hospital admission in patients with community-acquired pneumonia (CAP). By integrating XGBoost, LightGBM, and multivariable logistic regression—with post hoc calibration applied to the XGBoost model—the ensemble approach achieved superior predictive performance (AUC = 0.792), consistent calibration, and strong clinical decision-support capabilities. Additionally, interpretability was enhanced using SHapley Additive exPlanations (SHAP), which improved the model’s transparency and clinical applicability.

Based on an in-depth analysis of our findings, several key conclusions can be drawn. The ensemble model outperformed individual base learners in terms of both accuracy and robustness. By leveraging the complementary strengths of different algorithms and balancing feature-level biases, the weighted ensemble approach improved overall predictive capability. Respiratory rate emerged as the most important predictor of ARF in CAP patients, followed by the inflammatory markers tumor necrosis factor-α (TNF-α) and interleukin-1β (IL-1β). Elevated respiratory rate, along with increased levels of TNF-α and IL-1β, was positively associated with ARF risk. Other relevant predictors included heart rate, pleural effusion, and body temperature, whereas total bilirubin, serum calcium, albumin-to-globulin ratio (AGR), and platelet count contributed less to model prediction.

Our findings regarding the predictive importance of respiratory rate and heart rate are consistent with prior studies. In our cohort, patients who developed ARF had significantly higher respiratory rates (21.0 vs. 18.0 breaths/min, p < 0.001) and heart rates (103.5 vs. 91.0 bpm, p < 0.001) compared to those without ARF. Lyu et al. employed machine learning models to predict ARF in critically ill pneumonia patients in the ICU and similarly identified respiratory rate as a key feature [10]. Wang et al. developed a model for early ARF prediction in septic patients and also confirmed respiratory rate as an independent risk factor [11]. Elevated respiratory rate is often a compensatory response to hypoxemia, and a high rate at admission may serve as an early warning sign of oxygen deficiency. Bendavid et al. likewise found that both respiratory and heart rates were significant predictors of ARF in patients with COVID-19 [12].

In our study, TNF-α and IL-1β also emerged as critical inflammatory biomarkers associated with ARF risk. TNF-α is a potent pro-inflammatory cytokine that is activated early during infection and regulates a wide range of immune responses, including apoptosis, fever induction, and acute-phase protein synthesis. Lee et al. reported significantly higher TNF-α levels in both serum and bronchoalveolar lavage fluid (BALF) among ICU patients with fatal CAP compared to survivors, underscoring its association with disease severity and mortality [13]. Bacci et al. further validated these findings, noting elevated TNF-α concentrations in patients requiring mechanical ventilation, those with acute kidney injury (AKI), ICU admission, high CRB scores, and early mortality (<7 days) [14].

Similarly, IL-1β plays a central role in acute inflammatory responses and has shown promise as a prognostic biomarker. Secreted by macrophages, dendritic cells, and endothelial cells, IL-1β promotes immune cell recruitment and amplifies local inflammation through the stimulation of other cytokines. While Lee et al. did not observe significant differences in overall IL-1β levels between survivors and non-survivors, elevated BALF IL-1β concentrations were found in CAP patients with ARDS who did not survive, suggesting its role in localized pulmonary inflammation [13]. Bacci et al. also linked IL-1β with ARDS and early mortality, particularly in patients with high CRB scores [14]. Moreover, IL-1β has been associated with CAP-related complications such as pleural effusion and systemic inflammatory response syndrome (SIRS) [15]. A pediatric study also demonstrated that high IL-1β levels were associated with the need for pleural drainage in cases of pneumonia [16]. Collectively, these findings support the utility of IL-1β as a potential early biomarker for ARF risk stratification in CAP.

Pleural effusion has long been recognized as a critical factor influencing clinical outcomes in patients with community-acquired pneumonia (CAP). As a common complication of pneumonia, pleural effusion is associated with adverse outcomes, including prolonged hospital stays [18], increased risk of respiratory failure, and a higher likelihood of requiring mechanical ventilation [12,17–20]. It often reflects extensive pleural involvement, leading to lung compression and impaired expansion, thereby increasing the likelihood of ARF. In hypoxemic patients, pleural effusion can exacerbate oxygenation impairment. Notably, pleural effusion frequently precedes significant declines in oxygenation indices, providing a potential window for early risk identification.

Additional predictors identified in our analysis included body temperature, total bilirubin (TBIL), serum calcium, albumin-to-globulin ratio (AGR), and platelet count. Fever represents a host defense response that can enhance immune function [21]; however, excessive hyperthermia may adversely affect the cardiovascular, respiratory, and nervous systems and has been associated with increased mortality [22]. High fever (≥39.5°C) has been linked to poorer outcomes, likely due to increased metabolic demand, cardiovascular stress, and elevated oxygen consumption [23].

TBIL, a commonly used marker of liver function, also possesses antioxidant and anti-inflammatory properties. Nonetheless, elevated TBIL levels have been associated with worse outcomes in patients with sepsis, COVID-19, and other critical illnesses [24–26]. High TBIL may reflect impaired hepatic clearance, systemic inflammation, increased apoptosis, and oxidative stress, all of which can contribute to pulmonary injury and the development of ARF [27,28].

Abnormal serum calcium levels have been associated with poor prognosis and increased mortality in various conditions [29–33]. Hypocalcemia may lead to ARF through mechanisms such as muscle weakness, laryngospasm, or bronchospasm, whereas severe hypercalcemia can result in somnolence, confusion, or coma, ultimately precipitating respiratory failure [30].

Albumin and globulin, as major plasma proteins, reflect nutritional and immune status, respectively[34]. The albumin-to-globulin ratio (AGR) is considered a more robust prognostic indicator than either component alone. Abnormal AGR has been associated with unfavorable outcomes in several clinical contexts [35–37], and it may serve as an independent predictor of prognosis in both pneumonia and ARF [38,39].

Our analysis also revealed that elevated platelet counts were significantly associated with an increased risk of ARF. Excessive platelet activation can promote thrombosis, reduce pulmonary perfusion, and contribute to respiratory failure [40]. High platelet counts may reflect underlying systemic inflammation and coagulation imbalance, both of which are implicated in ARF pathogenesis. In CAP—particularly in pneumococcal infections—platelet activation has also been linked to cardiovascular complications such as myocardial infarction and stroke [41].

Despite the promising results, several limitations must be acknowledged. First, this was a retrospective, single-center study conducted at Beijing Chaoyang Hospital, which may introduce selection bias. The relatively small sample size (n = 257) and geographic limitation may reduce the generalizability of our findings. Second, the analysis was restricted to clinical data collected within the first 48 hours of admission, without longitudinal follow-up. As a result, dynamic changes in clinical status—such as treatment response or evolving vital signs—were not captured. Third, the model was only internally validated; external validation using larger, multi-center datasets is necessary to confirm its reliability and broader applicability.

## 6. Conclusion

This study developed a multi-model ensemble-based clinical prediction tool to evaluate the risk of acute respiratory failure (ARF) within 48 hours of hospital admission in patients with community-acquired pneumonia (CAP). Model interpretability based on SHAP (SHapley Additive exPlanations) highlighted the pivotal roles of clinical features such as respiratory rate, inflammatory markers (TNF-α and IL-1β), and heart rate in predicting ARF. The proposed model is well-suited for early risk stratification and personalized decision support in hospitalized CAP patients.

## 7. Declarations

Abbreviations: T, temperature; SBP, systolic blood pressure; DBP, diastolic blood pressure; Lac, lactate; AST, aspartate aminotransferase; ALT, alanine aminotransferase; LDH, lactate dehydrogenase; TG, triglycerides; TBIL, total bilirubin; DBIL, direct bilirubin; Alb, albumin; Glb, globulin; BUN, blood urea nitrogen; Na, serum sodium; K, serum potassium; Ca, serum calcium; P, serum phosphorus; Glu, glucose; Nt_BNP, N-terminal pro-brain natriuretic peptide; PT, prothrombin time; APTT, activated partial thromboplastin time; INR, international normalized ratio; WBC, white blood cell count; NEUT%, neutrophil percentage; RBC, red blood cell count; HCT, hematocrit; Hb, hemoglobin; Plt, platelet count; AG_Ratio, albumin-to-globulin ratio; CRP, C-reactive protein; PCT, procalcitonin; IL, interleukin; TNF, tumor necrosis factor; GCS, Glasgow Coma Scale.MLR:multivariable logistic regression model.LightGBM:Light Gradient Boosting Machine.Cal_XGBoost:calibrated Extreme Gradient Boosting.AUROC:area under the receiver operating characteristic curve.PPV: positive predictive value.NPV: negative predictive value.

## Ethics approval and consent to participate

The study has been approved by the Ethics Committee of Beijing Chaoyang Hospital, affiliated with Capital Medical University. Based on the committee’s review, the research uses only anonymized data and does not involve direct human participation. Therefore, the Ethics Committee has waived the requirement for written informed consent. The study adheres to the ethical principles outlined in the Declaration of Helsinki.

## Consent for publication

Not Applicable.

## Availability of data and materials

The datasets used and analysed during the current study available from the corresponding author on reasonable request.

## Competing Interests

No conflict of interest is declared by all the authors.

## Funding

Not Applicable.

## Authors’ contributions

YLX Contributed to data curation, conceptualization, data analysis, and manuscript writing. YC and QL Contributed to methodology and data curation. ADL Contributed to methodology. XM reviewed the initial draft. SBG Supervised and reviewed the manuscript. All authors approved the final manuscript and are responsible for its content.

## Acknowledgements

The authors would like to thank all those who contributed to the completion of this work.

## Notes

### Competing Interest Statement

The authors have declared no competing interest.

### Funding Statement

The author(s) received no specific funding for this work.

### Author Declarations

The study has been approved by the Ethics Committee of Beijing Chaoyang Hospital, affiliated with Capital Medical University. Based on the committee's review, the research uses only anonymized data and does not involve direct human participation. Therefore, the Ethics Committee has waived the requirement for written informed consent.

